# Climate Resilient Health Systems: A Case Study of Climate Change and Dengue in Nepal

**DOI:** 10.1101/2025.08.12.25333484

**Authors:** Mandira Lamichhane Dhimal, Bishal Dahal Khatri, Kjersti Mørkrid Blom-Bakke, Guna Niddhi Sharma, Pramod Joshi, Sailaja Ghimire, Ernst Kristian Rødland, Christopher D. Golden, Meghnath Dhimal

## Abstract

**Introduction:** Nepal is vulnerable to climate change and extreme weather events, resulting in the expansion of vector-borne diseases such as dengue throughout the country. Due to rising temperatures, dengue cases have been significantly increasing, underscoring the urgent need for a climate-smart approach to public health resilience. This case study assesses how the health system in Nepal has coped with the increase in dengue cases in relation to climate change over the last decade.

**Methodology:** We conducted a desk review concerning the impact of climate change on dengue in Nepal using WHO’s six-building blocks framework (leadership and governance, health workforce, health service delivery, health information system, and access to essential medicines). To complement the review, we also held two multi-stakeholder consultative workshops on dengue outbreak management convening academics, government (federal, provincial, municipal), the World Health Organization, private sector, and NGOs. One was conducted before the desk review and the other after the draft report was prepared to validate findings.

**Findings:** Our findings reveal that, despite comprehensive federal dengue policies and guidelines, implementation is undermined by limited local capacity, poor multisectoral collaboration, weak information transmission, and inadequate workforce and diagnostic resources. Strengthening health-worker and volunteer training, laboratory and surveillance systems (including collaborative, data-driven climate-smart public health platforms), meaningful community engagement, and sufficient resource allocation are essential to enable timely, locally tailored dengue prevention and control in Nepal.

**Conclusion:** Although national policies for dengue control are well established, local implementation suffers from limited capacity, poor coordination, and inadequate surveillance and diagnostics. Investing in workforce training, laboratory and data-driven surveillance enhancements, and community-centered engagement—backed by consistent resource allocation—is critical for timely, locally tailored dengue prevention and control in Nepal.

## Introduction

Nepal faces a triple planetary crisis, referring to the interconnected crises - climate change, pollution, and biodiversity loss. These three environmental challenges pose hazards to human well-being and ecological stability [1]. Nepal is experiencing different climate patterns and extreme weather events such as droughts, glacial melting, and a shift from snowfall to rainfall that leads to downstream floods and landslides and increasing wildfires, impacting a variety of sectors: agriculture, food and water security; human livelihoods; healthcare facilities and health system resilience; and air quality. In addition to these significant downstream effects, there is an expansion of vector-borne diseases such as dengue [2], which is attributed to climate change [3–5].

Changes in temperature, rainfall, and humidity influence the spread of dengue [6–8], in addition to unplanned urbanization, increased transportation, and inefficient mosquito control [9]. The spread by road, rail, and air transport aids geographical expansion of dengue in two ways: first, by movement pathways for viremic individuals (i.e., infected and asymptomatic), and second, by inadvertently transporting aedes eggs in tires and other cargo [10]. It is crucial to understand the patterns of disease mobility in the assessment of how climate change impacts dengue cases.

Most people with dengue have mild or no symptoms, but individuals who are infected a second time are at greater risk of severe dengue. There is currently no specific treatment for dengue; however, early detection and timely access to appropriate supportive care, such as fluid management and symptomatic relief—can significantly reduce mortality and improve outcomes. Elderly and children, mentally or physically challenged individuals, displaced individuals, and migrant or marginalized populations are disproportionately affected [11]. In Nepal, socio-economic factors such as illiteracy and poverty are associated with environmental conditions like exposure to stagnant water or containers where dengue-carrying mosquitoes are often found [12]. There is also an interplay between socioeconomic factors and access to information and healthcare utilization [13], partly due to high out-of-pocket payments and lack of health insurance [14], highlighting significant equity issues in the transmission of dengue.

The first dengue case in Nepal was reported in 2004 in a Japanese traveler to Nepal. The first dengue outbreak was in the southern Terai districts bordering India in 2006. Dengue and other vector-borne diseases have expanded from lowlands like Terai to higher areas like the Hindu Kush Himalayan region [15]. Nepal is now a dengue-endemic country. Dengue outbreaks in the last decade have overwhelmed the health system, which faces challenges in tackling both communicable and non-communicable diseases, among others due to overcrowded health facilities and resource constraints (human, financial, and logistics) [16].

Nepal faces a challenge in healthcare capacity to face this new issue. Nepal’s ratio of health professionals to its population is insufficient according to WHO standards [17]. The limited number of professional health care workers could lead to poor data collection of health information, under or over-utilization of medical equipment, and degradation in the quality of services [17]. The Ministry of Health and Population (MOHP) is the leading government agency responsible for the country’s health system, but both governmental and non-governmental organizations provide health services. The federal and provincial-level government oversees the specialized, secondary and provincial hospitals. The municipality level government is responsible for basic health service centers and primary hospitals. A significant gap exists between the availability of health workers in urban versus rural settings [17]. This is due to fewer incentives, improper accountability, and poor responsiveness of rural municipalities [17], in addition to scarce road infrastructure inadequate medical equipment, and few health facilities that also affect access to healthcare [14]. The decentralized health system, faces challenges due to limited resources and inadequate coordination with private entities and NGOs, and inter-ministerial and inter-sectoral collaboration, including overlapping roles [18]. In this case study, the overall objective is to assess how the health system in Nepal has coped with the increase in dengue cases in relation to climate change over the last decade.

## Methodology

### Stakeholder Consultative Workshops

Stakeholder consultation serves several critical functions in guiding research and policy development. By engaging those who are directly affected by—or who can influence—a given public health issue, consultation ensures that diverse perspectives and local knowledge inform every stage, from framing research questions to interpreting results [19]. This two-way dialogue helps to identify priorities that are both contextually relevant and actionable, fosters transparency around decision-making processes, and builds trust and ownership among participants [20]. We followed a four-step process[21]: First, we identified and prioritized stakeholders via a comprehensive desk review of policy documents, program reports, and published literature. Second, we provided context and rationale to the majority of stakeholders in an inception workshop. Third, we ensured ethical involvement by providing clear written information, emphasizing voluntary participation with withdrawal rights, focusing discussions on the topic rather than personal data, training facilitators on engagement best practices to avoid bias and power imbalances, and implementing confidentiality safeguards under ethics committee oversight. Finally, we structured our workshops into six breakout sessions aligned with WHO’s health-system “Six-building blocks” (service delivery, health workforce, information systems, access to essential medicines, financing, and leadership/governance) synthesizing information and without mentioning opinions of individuals. On 18 July 2024, we held the inception workshop (28 participants) to introduce the assignment and gather initial input. On 13 September 2024, a second workshop (31 participants) validated and supplemented the desk-review findings. Attendees included recovered dengue patients, university faculty, representatives of federal, provincial, and municipal governments, WHO, private organizations, and NGOs, ensuring broad perspectives. Fair compensation for time and expenses was provided to remove barriers and foster true dialogue, equitable contribution, and strong buy-in for the final document.

### Desk review

We conducted a qualitative thematic review of peer-reviewed articles, grey literature, government reports, and policy documents. We utilized the Reading, Extracting, Analyzing, and Distilling/reporting findings (READ) approach for systematic document analysis [22]. We used the following search databases for eligible peer-reviewed articles: Scopus, PubMed, Embase and Google Scholar, and we used the Google search engine for relevant grey literature and government reports. Policy documents were searched and downloaded from government websites. The retrieved information was extracted and organized.

We included articles from 2014 to 2024, focusing on Nepal; however, all the policy documents related to climate change and dengue were incorporated. Articles written in both Nepali and English language were included. We first searched articles by including key words like “Dengue” and “Nepal”, then we screened title and abstract, studies were included based on their relevance to the research question, methodology, and applicability of their findings. Duplicates and publications not addressing climate change, health systems, or dengue, were excluded from the review. The PRISM flow diagram was constructed from the Haddaway [23].

**Fig 1.**
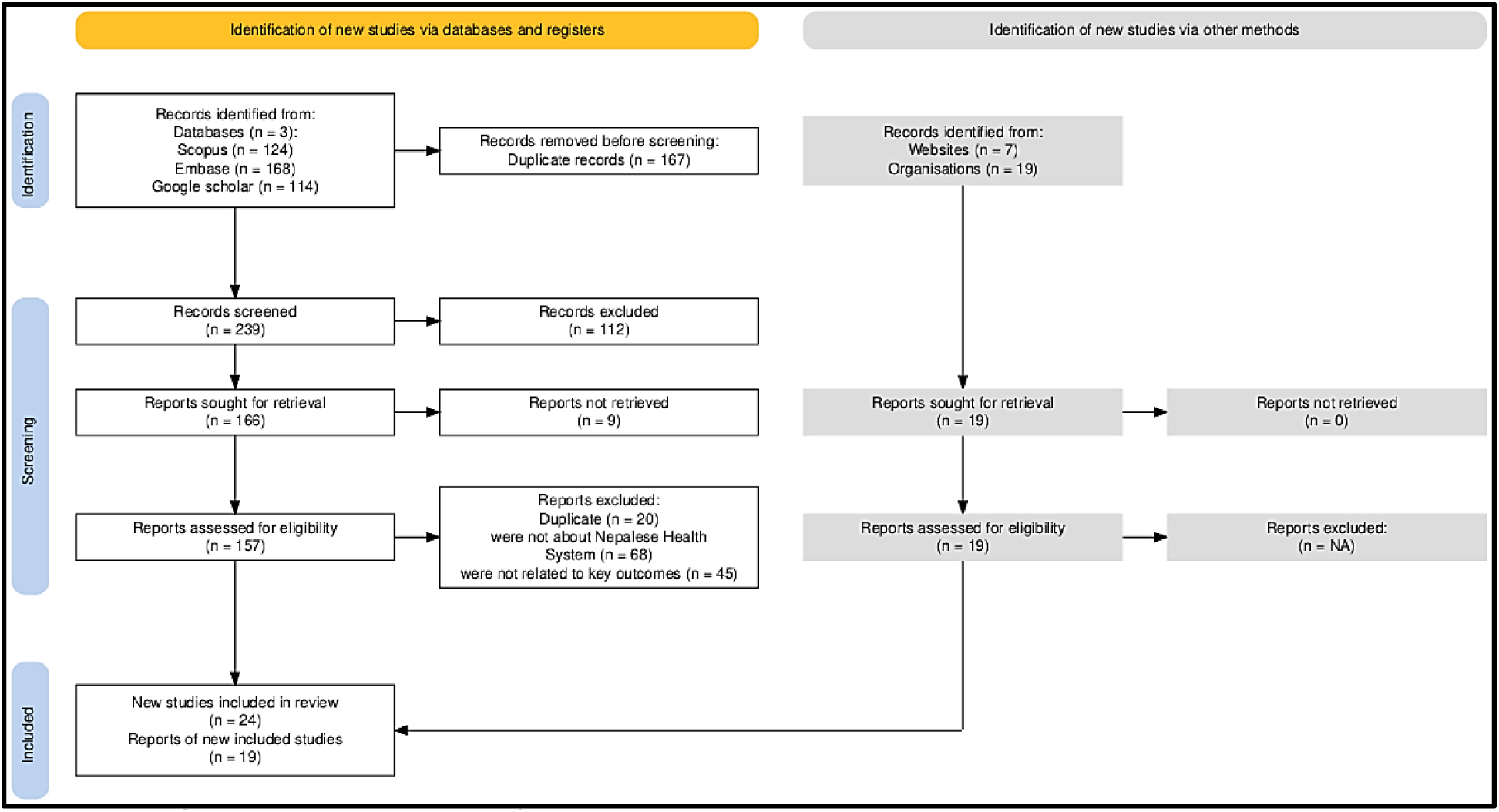
PRISM Flow Diagram of Literature search.

Data collection was guided based on the WHO Six-building blocks [24], reviewing each source for the following elements:

1. What policies are in place to address climate change’s impact on dengue in Nepal?
2. Who is involved in developing human resources for climate change and dengue?
3. How are dengue cases and climatic data reported in Nepal?
4. What technologies and measures are being applied to mitigate dengue?
5. What sort of climate and health programs are in place to prevent and control dengue outbreaks?
6. What kind of emergency preparedness plans are being deployed to mitigate dengue epidemic?
7. What are the sources of funding in Nepal for climate change and dengue? What is the status of this funding?
8. Any gaps, challenges or opportunities in mitigating the dengue epidemic in Nepal?

We conducted a thematic analysis following Braun and Clarke’s six-step method [25]: (1) familiarizing ourselves with the data, (2) generating initial codes, (3) searching for themes, (4) reviewing themes, (5) defining and naming themes, and (6) writing the final report. To structure our analysis, we also used a deductive approach based on the World Health Organization’s six health-system building blocks. These blocks informed both our criteria for the desk review and our coding framework for policy documents and stakeholder-consultation findings.

### Ethical approval

This study is primarily a desk review and conducted as a part of research study approved from the Ethical Review Board (ERB) of Nepal Health Research Council (Reg. No. 1977). In order to validate findings of desk review, stakeholder consultation workshop was carried out and feedbacks and suggestions are summarized without identifying individual level information. Before conduct of the workshop, oral consent of all workshop participants was taken which was approved by the ERB of Nepal as per national ethical guidelines for health research in Nepal.

## Findings

### Leadership and Governance - Policies and Partnership

#### Results from the Desk Review

Initiatives of dengue control were first introduced in the Three year Interim Constitution of Nepal (2007/08 to 2009/10) a bridge between the tenth and eleventh five-year plan [26], showcasing the commitment from the government towards dengue control. At present, MOHP is accountable for directing the dengue control program and ensuring coordination and resource allocation within the health system [27]. Several policy documents related to climate change and health exists (Table1).

**Table 1:**
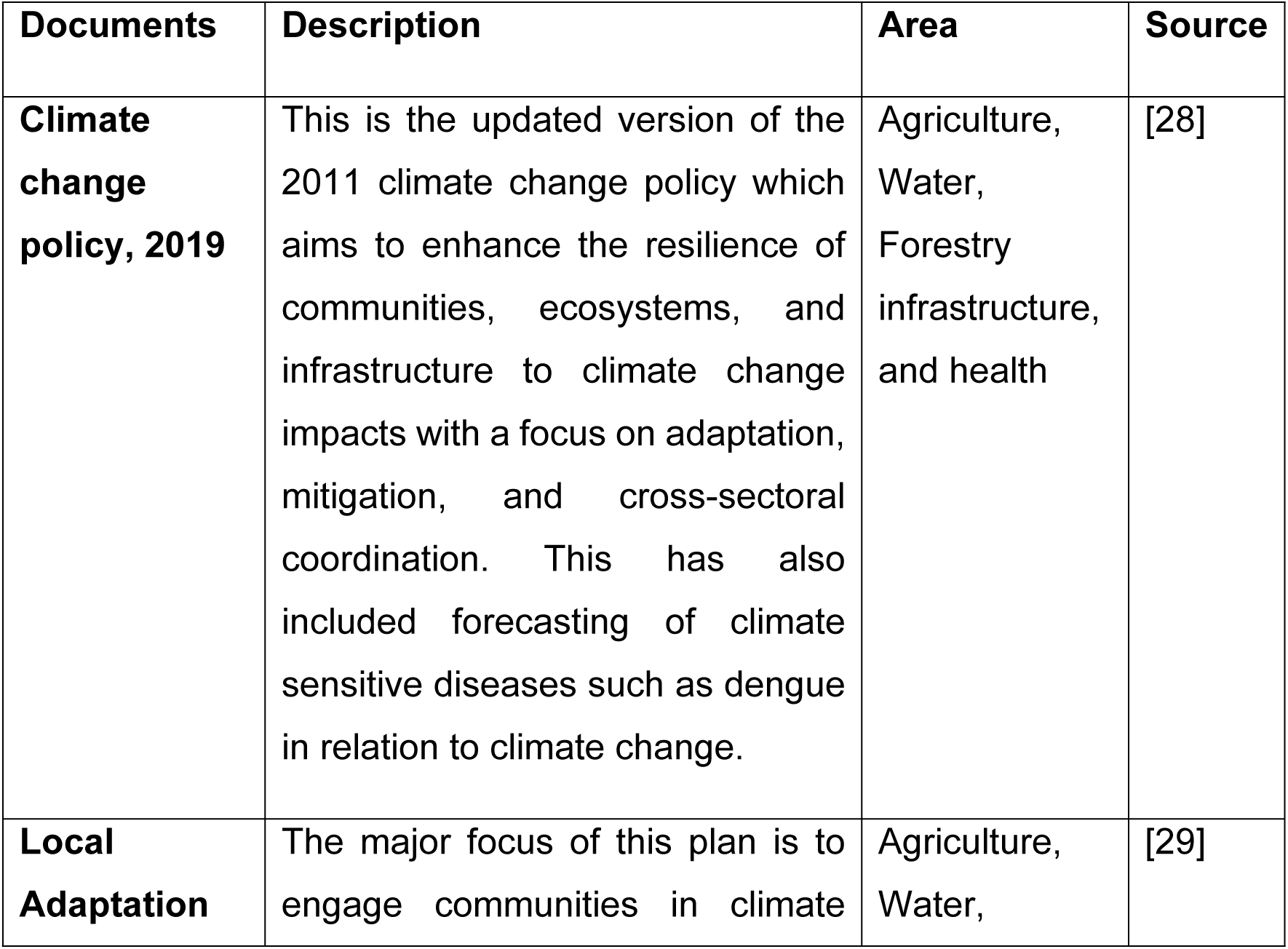
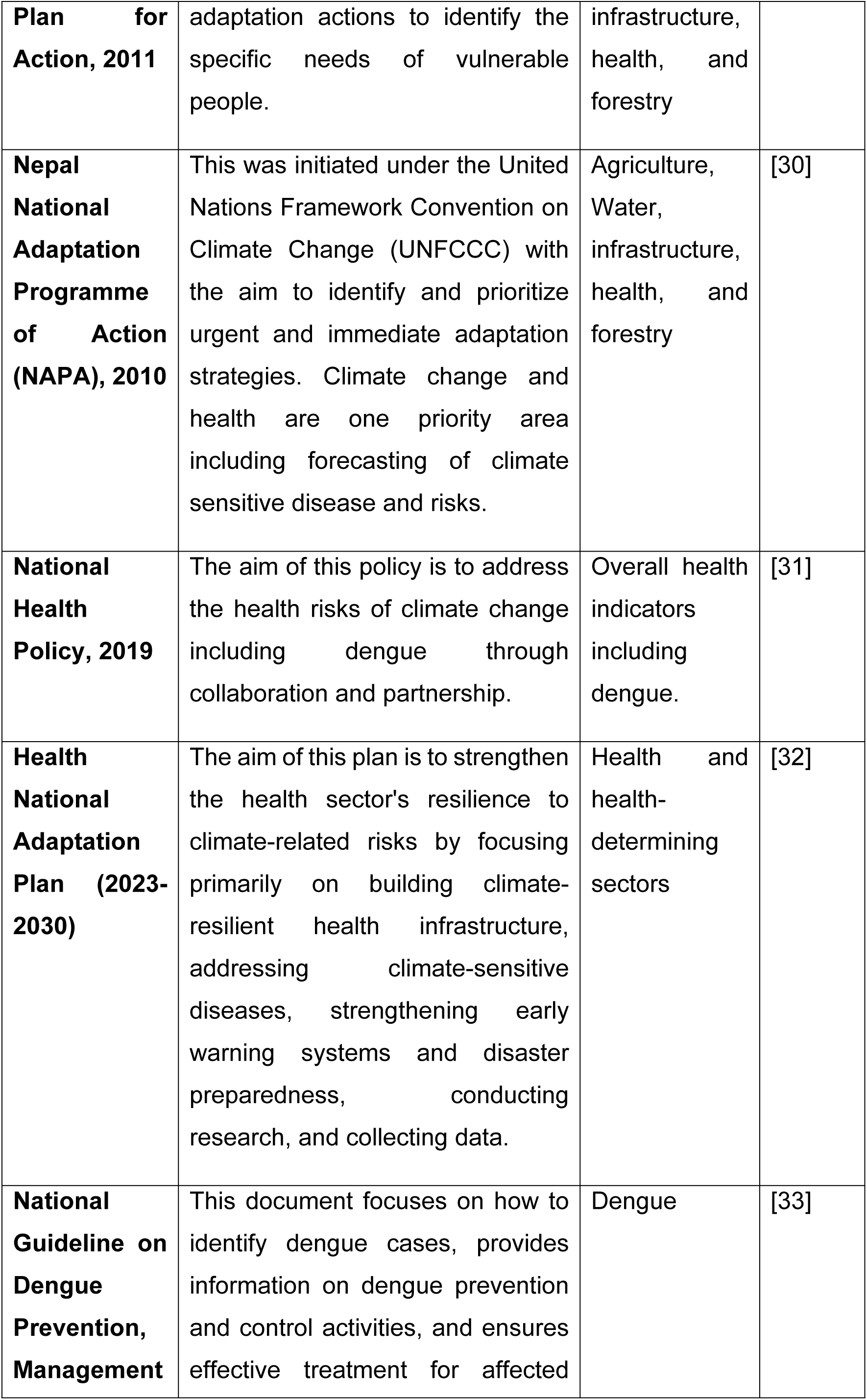
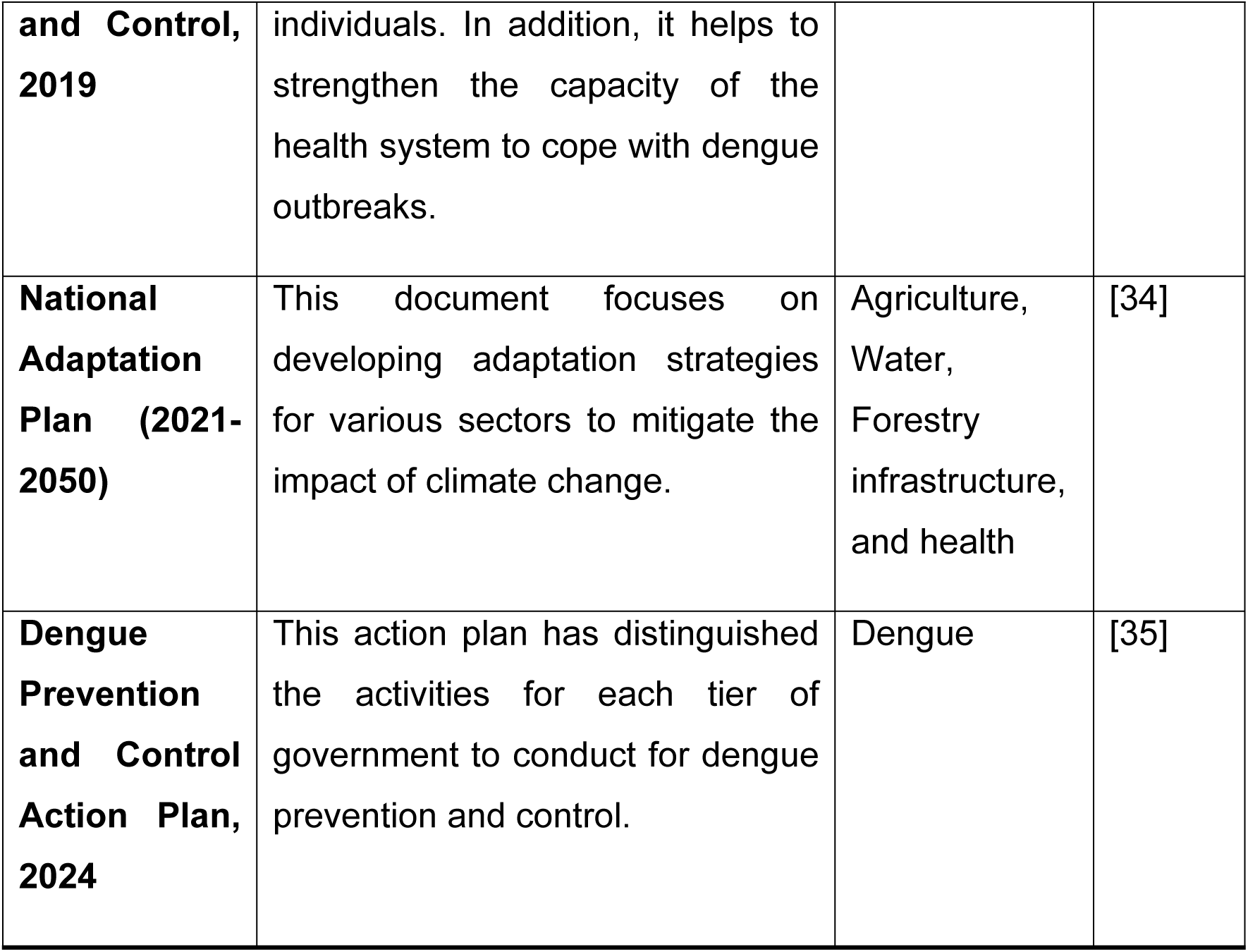
Policies and plans associated with climate change and dengue.

The national guideline on dengue prevention, management, and control, based on the 2008 WHO guideline, was updated in 2011 and 2019 to include internationally adopted definitions, protocols, and guidelines [33]. It is widely accepted by provincial and local governments. The 2024 MOHP action plan outlines the roles and responsibilities of each government tier and communities in dengue prevention, detailing activities for health system elements and non-health sectors [35].

Government bodies like the Epidemiology and Disease Control (EDCD) handle preparedness and outbreak response, including “search and destroy” campaigns to eliminate mosquito breeding sites [35]. The Nepal Health Research Council (NHRC), academic institutions, and NGOs contribute to evidence-based decision-making. The National Health Education, Information, and Communication Center develops materials to raise public awareness [36]. Nepal’s involvement in the regional technical advisory group on dengue shows its commitment to control efforts [18]. Local government run various dengue control programs.

Despite existing guidelines, challenges such as ineffective monitoring, inconsistent implementation, and weak surveillance hinder accurate dengue risk assessment. A robust dengue response plan at the municipality level is needed to mitigate future outbreaks”[37].

#### Results from the Stakeholder Consultations

Some local government actors have developed their own standard operating procedures for search and destroy of *Aedes* mosquitoes breeding places, while others follow the standard operating procedures developed by the EDCD. In some local government, mayors have shown interest in dengue management, prevention, and control, requesting situation updates on dengue cases.

Chandragiri Municipality demonstrated strong local leadership and multisectoral collaboration in dengue prevention establishing dedicated response committees, partnering closely with the Health Office, and empowering ward focal persons while engaging community volunteers and professionals through targeted education, conducting “search and destroy” field operations and awareness campaigns, and allocating NPR 2.5 million to vector control in the fiscal year 2080/81 BS, outreach, and healthcare preparedness.

Despite having documents and necessary information on dengue control and prevention, most local leaders are unaware of this information due to inadequate flow of information.

### Health workforce

#### Result From the Desk Review

Dengue prevention and control requires a holistic approach, and needs diverse health professionals such as entomologists, epidemiologists, public health professionals, community health workers, physicians, and clinicians[33]. Several studies indicate that there is an insufficient number of health workers as dengue is rapidly expanding its geographical range [17,18,38]. Frequent transfer of health professionals, and high staff turnover rate are additional challenges for managing human resources [18]. Nepalese health institutions provide certain allowances for health workers working in high-risk zones, but further policy actions are still needed.

Capacity building of clinicians in the management of dengue cases is needed based on the distribution and burden of dengue cases [37]. Capacity-building activities are being carried out such as clinical seminars, and integrated trainings [18]. The Government of Nepal has provided tire waste management training to auto mechanic associations, recollections, and recycling associations. [18] Furthermore, they have educated journalists and media to disseminate information to the public. [18] A malaria-dengue taskforce was also created. [39] Even though workforce development activities have been executed, the number of skilled health workers remains inadequate and there are few refresher training programs. A significant number of health workers are unaware of the dengue reporting system, and are only able to report symptomatic cases due to lack of skills and diagnostics, which hampers the quality of data [40].

#### Results from the Stakeholder Consultations

The National Health Training Center train additional health professionals and has initiated training in the use of the Surveillance Outbreak Response Management and Analysis System (SORMAS). Capacity building of health professionals and medical recorders working at federal, provincial and local governments is still needed, although orientation activities on dengue control and prevention have been conducted in past years, frequent staff movements and insufficient skilled health personnel in dengue area further hinders enhancing capacity building. Other challenges are the lack of refresher training, coupled with a high turnover rate among health personnel.

Entomological labs were recently built in each of the 7 provinces where 20-30 staff members per province received training on vector control. Without any financial incentives or daily allowances, the tendency of staff to take up entomology work has diminished, as one must put in morning and evening shifts to accomplish the entomological work (outside official working hours). Secondly, there are no separate posts for entomologists in these laboratories, which further complicates the matter. While the number of competent vector control supervisors who have specialized training in mosquito control is high, they are not being placed in appropriate positions. Both on the federal and provincial levels, posts for entomologists are either not available or filled by persons with other expertise.

### Health Information System - Data and surveillance

#### Results from the Desk Review

Dengue cases are reported through the Early Warning Reporting System (EWARS) from sentinel sites using District Health Information Software (DHIS2), and SORMAS in some provinces [18]. The situation of dengue is updated by the EDCD [40]. There is a reporting loop from the 118 sentinel sites to the Vector-Borne Diseases Research and Training Center, to EDCD that provides feedback to the district level. Data on vector distribution is crucial to predict the outbreak of vector-borne diseases, even though EWARS has not considered the integration of vector surveillance in their system [41].

The EWARS does not provide any specific guidelines or a numeric threshold for an outbreak. Criteria must be established to define an outbreak to systematically identify when an outbreak is likely to occur [16]. Alarm signals can be developed based on meteorological or entomological indicators as done by Singapore, Malaysia, and other dengue-endemic countries [16]. Climatic data (e.g., temperature, rainfall) are available from meteorological stations located all over Nepal, and these data can be used to predict meteorological changes. Alteration in rainfall, humidity, and temperature is linked with heightened dengue cases [3–5]. Thus, it is possible to anticipate at what time of the year or month the risk of dengue outbreaks or other vector-borne diseases are likely to occur.

Innovations, such as geographic information systems (GIS) remote sensing, digital tools, and the use of mobile applications such as Nepadengue [42] offer an opportunity to assess the risk of dengue in the future. Nepadengue was developed to assist the health system by reporting breeding sites and taking action accordingly [42]. However, the application is yet to be tested in the broader context of the Nepalese health system. The limitation included digital literacy gaps, connectivity constraints, and device availability, while the future potential could be government ownership, usability enhancement, and broader piloting [42]. To date, climatic variables are not used to develop forecasting of dengue and an early warning system in Nepal but it’s in progress in collaboration with World Health Organization and University of Oslo.

#### Results from the Stakeholder Consultations

Stakeholders noted a critical lack of research on how climatic and non-climatic factors (including vector biology, infectivity, and co-infections with Zika or chikungunya) influence dengue in Nepal, and pointed out that no studies have quantified climate change’s attribution to dengue burden. Until 2019, irregular reporting and low prioritization hindered accurate risk estimation, even though an outbreak is officially defined as over 300 cases in a cluster or more than two standard deviations above normal. The EWARS System provides weekly dengue data but misses cases outside sentinel sites and varies in its inclusion of inpatient versus outpatient cases, creating inconsistent surveillance. Local authorities rarely know exact mosquito breeding locations and resources are often deployed haphazardly, while planned hotspot mapping remains incomplete. To address these gaps, stakeholders recommend a unified, efficient reporting platform—potentially via mobile health apps—and the integration of high-resolution climate and epidemiological data to enable real-time, precise risk models. Promising steps include the new Integrated Health Information Management System (IHIMS) upgrade linking climate with health data, line-listing of dengue cases, scaling up SORMAS, and piloting a Climate and Health Analytical Platform using DHIS-2 data to deliver an early warning system adaptable across the country.

### Health technologies, medical products, and supply chain

#### Findings from the Desk Review

Methods like PCR and ELISA are required to diagnose dengue cases, and it is possible by means of rapid diagnostic kits crucial for detecting dengue in the early stage and providing prompt treatment [43]. The EDCD, and international organizations like WHO have provided rapid diagnostic kits and logistic support to various health facilities and the National Public Health Laboratory [44]. However, there is frequent shortages of rapid diagnostic kits during epidemics. Even though health facilities are equipped with the necessary tools, reliable confirmation of the dengue virus remains a challenge in rural areas. Disparities in resources and capacity between local government often lead to inconsistent service delivery, exacerbating the difficulties in diagnosing dengue accurately in rural settings [38]. Poor supply chain logistics also hinder the availability of medical equipment [17].

#### Findings from the Stakeholder Consultations

Introducing new technologies like controlling dengue vectors through biological methods and installing vector sensing equipment could be a major milestone towards preventing and controlling dengue. Stakeholders stressed the need for entomological labs in all 77 districts along with comprehensive surveillance systems to monitor dengue infection and its vector.

### Service Delivery

#### Findings from the Desk Review

Activities to modify and manipulate environmental determinants are taking place in Nepal, such as utilizing led-on water storage, clean-up campaigns, search and destroy vector campaigns, waste management to reduce breeding places, and covering or destroying sources of stagnant water. Furthermore, there are guidelines related to water quality monitoring [45]. The introduction of smart agriculture has reduced the vector density owing to well-managed water inputs [46].

The Government of Nepal has launched a vector control program (search and destroy), an awareness program, and an early case detection program to prevent and control dengue [40]. Education, awareness campaigns, and community engagement are implemented to change human behavior[33]. EDCD and the National Health Education, Information, and Communication Center are delivering awareness programs to the community using information, education, and communication materials [36]. EDCD has organized numerous review meetings with diverse stakeholders on the status of dengue preparedness and strategies to mitigate them, and is working accordingly. [44]. The Government of Nepal has provided health information through diverse sources, such as audio sources (radio, mobile phone), audio-visual platforms (Facebook, TVs), and visual materials (posters, and pamphlets) in collaboration with the National Health Education, Information, and Communication Center [36]. Digital interaction and communication regarding dengue prevention and control between EDCD, local government, district, and province is ongoing to halt dengue transmission. [47]. The Health Emergency Operation Centre is the main government body that looks after public health-related emergencies such as dengue outbreaks.

Although awareness activities are being carried out, people are still unaware of how to prevent and control dengue. A study conducted by Bhattarai and colleagues highlighted that the knowledge regarding dengue prevention and control among surveyed participants were found to be moderate (with mean score of 54.20) [48–51]. Further, despite efforts by the government, unplanned and unmanaged urbanization poses the greatest threat to dengue transmission as it leads to unmanaged drainage, and crowded and congested places that could be potential breeding sites for dengue vectors [41]. Inter- and intra-sectoral coordination such as with transport, WASH, and coordination among different tiers of government is needed for more concerted efforts [33].

#### Results from the Stakeholder Consultations

Stakeholders discussed several possible preventive measures for preventing and controlling dengue. For example, they mentioned the use of Bacillus thuringiensis (Bti) which is a naturally occurring bacteria used as a control measure to combat the dengue larvae. Additionally, they mentioned spraying insecticides to kill adult dengue vectors and targeted indoor residual spraying, which is an insecticide sprayed in the walls and ceilings of houses to kill dengue vectors. They further discussed that these measures are often implemented without proper technical assistance or adherence to established guidelines. This uncoordinated approach raises concerns about the potential development of insecticide resistance, which could complicate future control efforts including adverse impacts on human and environment. The main aim of these health programs is to reduce morbidity due to dengue and achieve the target of zero deaths. However, ambiguity persists among local leaders and health professionals as to how to effectively carry out activities for the control of dengue. Participants shared that spraying insecticides has been reduced compared to previous years, while stakeholders stressed that if dengue is not controlled after search and destroy campaigns, they have no alternative options other than to use fogging.

In addition, hygienic measures have been implemented. For example, environmental cleanliness activities to destroy mosquito breeding sites are conducted every Friday on the premises of health facilities, such as health posts and health offices. Other activities include raising awareness through mobile phone ringtones and sharing information, education, and communication, and conducting behavioral change communication programs. Dengue testing in the community has increased owing to dengue cases registers available at the primary health care center level. This has resulted in earlier detection and response, and improved the accuracy of reporting and recording of cases.

A program conducted by the Disaster Response Emergency Fund from June to December 2023 to prevent and control the spread of dengue in Kathmandu Valley (Kathmandu, Bhaktapur, and Lalitpur districts) was scaled up in 13 additional districts (about 2 million people) in coordination with local Red Cross and Red Crescent members. Strategies included strengthening health systems, involvement of community members in raising awareness, assisting search and destroy campaigns, and disseminating information were executed to reduce dengue risk, in addition to an awareness program on Water Sanitation and Hygiene (WASH). The program was viewed as successful. However, the development of information, education, and communication materials in the local language, and early community engagement could yield even better results. Thus, this elucidates the importance of community engagement and awareness campaigns in reducing dengue in Nepal.

Strengthening community participation is a key to reduce the impact of future outbreaks. Most of the stakeholders highlighted that door-to-door awareness about dengue is required since most citizens do not have the necessary knowledge of ways to prevent and control this disease. In the future, health programs should focus on integrated vector management and community engagement to ensure that they are successful. Stakeholders also proposed the principle of community-based leadership to safeguard that service delivery is done on time.

### Financing

#### Results from Desk Review

The Nepal budget speech of 2021/23, included funding for search and destroy campaigns for 139 local government [47]. Investment in other sectors such as WASH and climate change indirectly helps to prevent and control dengue as there has been a rise in funding for these relevant sectors [52]. Furthermore, expectations from communities are high for mosquito-repellent instruments [53]. Nepal’s inability to address inequalities and passive purchasing poses a major threat to funding for dengue control [54].

#### Results from Stakeholder Consultation

The Government of Nepal has separated a conditional grant of NRs 100million from its total budget in the fiscal year 2023/2024 and has distributed those funds as per the number of cases taking the dengue burden (incidence) of the last three years. 420 out of 753 local government have received the sum of NR 79.66million ranging from NR 50 thousand to NR 8 hundred thousand and each of the 77 districts NR 200 thousand. Furthermore, local government have emergency funds that they can utilize during disease outbreaks, such as dengue.

Although there is provision of funding, its efficient and effective utilization to prevent and manage dengue is still a question due to the lack of monitoring and evaluation tools for dengue control and prevention. In addition, stakeholders stressed that there is limited funding from WHO, community-based organizations, Red Cross, and other development partners for dengue control and prevention. There is a need to increase both domestic and international funding for addressing emerging health risks of climate change such as dengue.

## Discussion

Our study addresses how the Nepalese health system has coped with the rising dengue cases in relation to climate change over the last decade. This study centers around the six building blocks of WHO [24], highlighting the linkage between health system, dengue and climate change in Nepal.

Under the theme of leadership and governance, our study found that Nepal has established several key policies, guidelines, and action plans at the central level to tackle dengue, reflecting strong governmental commitment to vector-borne disease control [55]. In some forward-thinking local government with engaged local leaders and communities, these policies have been translated into integrated public health prevention and control measures. However, implementation is hampered by limited capacity, insufficient multidisciplinary collaboration, and fragmented information flows across the central, provincial, and local tiers of government [16]. To strengthen Nepal’s response, especially as climate change expands the ecological niche of *Aedes* mosquitoes, national guidelines should be aligned with the World Health Organization’s 2024 *Aedes*-borne Disease Global Strategic Preparedness and Response Plan, incorporating international best practices to ensure both local effectiveness and global conformity [55]. Finally, formalizing intersectoral coordination committees with clear communication protocols and adopting cloud-based platforms with role-based access control would facilitate timely sharing of entomological, clinical, and epidemiological data among public health, clinical, and vector-control teams, thereby closing critical gaps in information exchange and collaboration [56].

Similar to past research [58], our study reveals that shortages of trained health and vector-control personnel, especially during peak dengue seasons, undermine clinical management, surveillance, and timely reporting. Knowledge gaps among health workers and communities further impede source reduction and early case detection [57]. To strengthen Nepal’s workforce, retention incentives and performance-based rewards, plus interdisciplinary training that brings together entomologists, health educators, and waste-management officers should be prioritized.

Our study highlighted that community engagement that includes meaningful and actionable strategies, tailored to the local context and across sectors can prevent transmission and supports control efforts. Enhancing community capacity and informing them on dengue prevention and control should be expanded with culturally tailored Information, Education and Communication materials co-developed by local leaders [58]. Stakeholder participation and local ownership are vital elements for sustainable implementation of dengue prevention and control measures. Finally, addressing waste-management and insecticide resistance, formalizing cross-border surveillance with India, and scaling volunteer community-agent networks will bolster Nepal’s dengue resilience [16,41].

Our study highlights that Nepal’s reliance on passive, hospital-based syndromic reporting with limited laboratory confirmation leads to delayed and under-reported dengue case detection, hampering timely response [16,41]. Integrating meteorological data like temperature, rainfall, and humidity—into DHIS-2 can strengthen outbreak prediction, as demonstrated by smartphone fever reporting and GIS mapping trials elsewhere [59]. However, scale-up is constrained by technical capacity, resource shortages, and fragmented data-sharing protocols [16,41]. Aligning with the WHO’s 2024 Global Strategic Preparedness and Response Plan, a multi-pronged surveillance approach: augment traditional reporting with climate-informed predictive models, sentinel-site surveillance in newly affected districts, and clear data-sharing agreements among health, meteorological, and academic stakeholders could be essential to mitigate dengue [55]. Employing and piloting mHealth tools like “NepaDengue” [42] for community-level reporting can further close surveillance gaps. By combining passive, active, and predictive surveillance streams, Nepal can build a timely, flexible early-warning system that optimizes dengue prevention and control in a changing climate.

Our study confirms that weak diagnostic capacity characterized by recurrent stock-outs of ELISA kits and PCR reagents undermines timely dengue detection and surveillance, particularly in climate-risk areas where laboratory and cold-chain infrastructure are often interrupted. To address these gaps, decentralizing diagnostics through serotype NS1 rapid tests that require minimal infrastructure and training, and deploying solar-powered isothermal amplification platforms for in-field molecular confirmation in settings lacking conventional labs can be implemented [60]. Strengthening laboratory staff capacity and ensuring reliable access to quality rapid diagnostic kits will be critical for outbreak response. Furthermore, monsoonal floods and landslides frequently disrupt supply chains for diagnostics, therapeutics, and vector-control commodities [61].

Under the theme of financing, our study found that although policymakers and health system leaders strive to allocate resources amid competing demands, current funding remains insufficient to support dengue prevention and control across all local government. In low-income settings like Nepal, dengue outbreaks impose substantial out-of-pocket costs on affected households, exacerbating economic hardship despite limited local data [62]. Constrained health budgets and competing priorities often leave vector-borne disease programs underfunded, perpetuating deficits in leadership, workforce capacity, and access to critical technologies. To address this, embedding specific financial modalities for climate-adapted dengue control within broader health and development plans will be essential for equitable, responsive resourcing and for safeguarding vulnerable communities and the overburdened health system [63].

One of the main limitations of our study is that we were only able to carry out systematic review of literature. Additionally, we relied on a single, one-day workshop for stakeholder input whereas a longer, more in-depth series of meetings or analyses would likely have uncovered richer insights. These constraints were driven by the resources available to our team.

## Conclusion

Despite robust federal level policies and guidelines targeting dengue prevention, effective implementation at the local level remains constrained by insufficient technical capacity, fragmented multidisciplinary coordination, and delayed information flow across government tiers. Critical gaps in workforce training, diagnostic infrastructure, and real-time surveillance undermine timely outbreak responses. Strengthening these areas—through targeted capacity-building for health professionals and volunteers, enhancement of laboratory and collaborative surveillance systems, and sustained resource allocation alongside meaningful, context-specific community engagement and stakeholder ownership, is essential to translate policy into practice and achieve sustainable dengue control in Nepal.

## Data Availability

This article does not report any primary data. All findings are based on a desk review of published literature and stakeholder consultations, during which no individual level or sensitive data were collected.

## Funding

The seed funding for this case study was provided by Norwegian Agency for International Development (NORAD). This research study is also linked to the research project entitled “Health Policy and System Research Responding to Climate Crisis: a Lesson from Nepal” funded by the Alliance for Health Policy and System Research, World Health Organization and “Big Data Climate-Health Analyses Across Infectious and Non-Communicable Diseases and Nutritional Disorders in Nepal” funded from Harvard T. Chan School of Public Health. There is no role of funding body on research design and interpretation of findings. The views expressed in this case study are solely those of the authors and do not reflect the views of any affiliated institutions.

## Acknowledgements

We are thankful to Dr. Idil Shekh Mohamed and Dr Robert Marteen of the Alliance for Health Policy and System Research, Science Division, World Health Organization, Geneva, Switzerland for their constructive feedback and suggestions to prepare this manuscript.

## Notes

### Competing Interest Statement

The authors have declared no competing interest.

### Clinical Trial

NA

